# Outcomes of hydroxychloroquine usage in United States veterans hospitalized with Covid-19

**DOI:** 10.1101/2020.04.16.20065920

**Authors:** Joseph Magagnoli, Siddharth Narendran, Felipe Pereira, Tammy Cummings, James W. Hardin, S. Scott Sutton, Jayakrishna Ambati

**Affiliations:** Dorn Research Institute, Columbia VA Health Care System, Columbia, South Carolina, USA; Department of Clinical Pharmacy & Outcomes Sciences, College of Pharmacy, University of South Carolina, Columbia, South Carolina, USA; Department of Epidemiology & Biostatistics, University of South Carolina, Columbia, South Carolina, USA; Center for Advanced Vision Science, University of Virginia School of Medicine, Charlottesville, Virginia, USA; Department of Ophthalmology, University of Virginia School of Medicine, Charlottesville, Virginia, USA; Department of Pathology, University of Virginia School of Medicine, Charlottesville, Virginia, USA; Department of Microbiology, Immunology, and Cancer Biology, University of Virginia School of Medicine, Charlottesville, Virginia, USA

## Abstract

**BACKGROUND:** Despite limited and conflicting data on the use of hydroxychloroquine in patients with Covid-19, the U.S. Food and Drug Administration has authorized the emergency use of this drug when clinical trials are unavailable or infeasible. Hydroxychloroquine, alone or in combination with azithromycin, is being widely used in Covid-19 therapy based on anecdotal and limited observational evidence.

**METHODS:** We performed a retrospective analysis of data from patients hospitalized with confirmed SARS-CoV-2 infection in all United States Veterans Health Administration medical centers until April 11, 2020. Patients were categorized based on their exposure to hydroxychloroquine alone (HC) or with azithromycin (HC+AZ) as treatments in addition to standard supportive management for Covid-19. The two primary outcomes were death and the need for mechanical ventilation. We determined the association between treatment and the primary outcomes using competing risk hazard regression adjusting for clinical characteristics via propensity scores. Discharge and death were taken into account as competing risks and subdistribution hazard ratios are presented.

**RESULTS:** A total of 368 patients were evaluated (HC, n=97; HC+AZ, n=113; no HC, n=158). Rates of death in the HC, HC+AZ, and no HC groups were 27.8%, 22.1%, 11.4%, respectively. Rates of ventilation in the HC, HC+AZ, and no HC groups were 13.3%, 6.9%, 14.1%, respectively. Compared to the no HC group, the risk of death from any cause was higher in the HC group (adjusted hazard ratio, 2.61; 95% CI, 1.10 to 6.17; P=0.03) but not in the HC+AZ group (adjusted hazard ratio, 1.14; 95% CI, 0.56 to 2.32; P=0.72). The risk of ventilation was similar in the HC group (adjusted hazard ratio, 1.43; 95% CI, 0.53 to 3.79; P=0.48) and in the HC+AZ group (adjusted hazard ratio, 0.43; 95% CI, 0.16 to 1.12; P=0.09), compared to the no HC group.

**CONCLUSIONS:** In this study, we found no evidence that use of hydroxychloroquine, either with or without azithromycin, reduced the risk of mechanical ventilation in patients hospitalized with Covid-19. An association of increased overall mortality was identified in patients treated with hydroxychloroquine alone. These findings highlight the importance of awaiting the results of ongoing prospective, randomized, controlled studies before widespread adoption of these drugs.

---

The rapidity of the Covid-19 pandemic has exerted inordinate pressure on clinicians and drug regulatory agencies throughout the world to expedite development, approval, and deployment of both experimental drugs and repurposing of existing therapeutics. Among the myriad therapeutics advanced as potential repurposing candidates for Covid-19, the antimalarial and immunomodulatory drug hydroxychloroquine has captured great attention following an open-label, non-randomized, single treatment center study that reported efficacy of hydroxychloroquine and a potential synergistic effect with the macrolide antibiotic azithromycin, in improving viral clearance in Covid-19 patients.^1^ The resulting spotlight and public interest has led to its soaring utilization in Covid-19, drug shortages impacting its use in labeled indications, and stockpiling by countries.

Subsequent studies have not identified a similar benefit of hydroxychloroquine in Covid-19^2-5^ and concerns have been raised about the original positive study.^6,7^ Nevertheless, the United States Food and Drug Administration used its emergency authority for only the second time ever to permit the use of a drug for an unapproved indication^8^ in the case of hydroxychloroquine for Covid-19 in situations where clinical trials are unavailable or infeasible.^9^

Multiple prospective, randomized trials of hydroxychloroquine are now underway and will, in due course, provide valuable information about safety and efficacy. However, given its increasingly widespread use, not only as therapy but also as prophylaxis for Covid-19, there is a great and immediate need to obtain insights into the clinical outcomes among patients currently treated with hydroxychloroquine, particularly because of the non-negligible toxicities associated with its use.

Therefore, we conducted a retrospective analysis of patients hospitalized with Covid-19 in all the Veterans Health Administration medical centers across the United States to analyze the associations between hydroxychloroquine and azithromycin use and clinical outcomes. The findings of this nationwide study of one of the most complete national datasets in the United States can accelerate our understanding of the outcomes of these drugs in Covid-19 while we await the results of the ongoing prospective trials.

## METHODS

### STUDY DESIGN

This national retrospective cohort study evaluated information on hospitalized patients with confirmed SARS-CoV-2 infection using data from the Department of Veterans Affairs (VA). Data were extracted from the Veterans Affairs Informatics and Computing Infrastructure (VINCI), which includes inpatient, outpatient data (coded with International Classification of diseases (ICD) revision 9-CM, revision 10-CM), laboratory, and pharmacy claims. The completeness, utility, accuracy, validity, and access methods are described on the VA website, http://www.virec.research.va.gov. This study was conducted in compliance with the Department of Veterans Affairs requirements, received VA Institutional Review Board, and VA Research & Development approval.

### STUDY POPULATION

We developed a cohort comprising patients with laboratory confirmed SARS-CoV-2 infection in an inpatient setting. SARS-CoV-2 status was classified by laboratory results that were extracted from VA laboratory data. A text search for SARS-CoV-2 laboratory tests was used to query VA lab results. The study index date was defined as the date of a hospitalization with a positive SARS-CoV-2 laboratory test. Index dates range from March 9, 2020 to April 11, 2020, and patients were followed from index until hospital discharge or death. The period prior to index is designated as the baseline period and on or after index is designated the follow-up period. Patients were included in the study if their information included 1) a body mass index, 2) vital signs during an encounter (temperature, heart rate and blood pressure), and 3) discharge disposition status available for the hospitalization.

### OUTCOMES AND EXPOSURE CODING

The study outcomes are the result of the hospitalization (discharge or death), whether ventilation was required, and the result of hospitalization among patients requiring ventilation. Ventilator usage was coded using HCPCS/CPT codes (31500, 94003, 94002, E0463) and ICD-10-PCS codes (5A0955, 5A0945, 5A0935, 5A1522F, 5A1522G, 5A1522H). The results of the hospitalization were coded from the discharge disposition status on the inpatient record. Hospitalization data were taken from the VA inpatient hospitalization data.

Patients were assigned to one of three cohorts based on medication exposure to hydroxychloroquine (HC) and azithromycin (AZ): 1) HC-treated; 2) HC- and AZ-treated; or 3) HC-untreated. Patients were exposed to hydroxychloroquine if they had a dispensed drug from the VA bar code medication administration (BCMA) data file during their hospitalization. Similarly, if patients received azithromycin with hydroxychloroquine during their hospitalization they were categorized HC- and AZ-treated. Patients with no hydroxychloroquine exposure were coded as HC unexposed. To examine the association with ventilation, the time of hydroxychloroquine and azithromycin dispense was coded dynamically, before or after ventilator support.

### COVARIATES

At baseline (date of admission), for each patient, we extracted demographic, comorbid, clinical (vital sign) and pharmacy data including information associated with increasing severity of Covid-19.^10,11^ Demographic and clinical characteristics included age, sex, race, and body mass index (BMI). For comorbid conditions, we utilized ICD-10-CM codes and calculated the Charlson comorbidity index from relevant patient data. Vital sign data include heart rate, pulse oximetry, respirations, temperature, and blood pressure (BP). All vital sign data were collected at the first set of vital results during the patient’s hospitalization and all were prior to ventilation if applicable. Laboratory data during hospitalization were also evaluated for each patient and consisted of liver function tests, albumin, bilirubin, creatinine, blood urea nitrogen, erythrocytes, hematocrit, platelets, white blood cells, C-reactive protein, procalcitonin, troponin, and erythrocyte sedimentation rate.

### STATISTICAL ANALYSIS

The statistical analysis for this study was conducted in multiple steps. First, we generated summaries of the baseline demographic, comorbid, and clinical characteristics for each cohort treatment group (HC, HC+AZ, and no HC). To summarize differences across treatment groups, continuous variables were analyzed with the ANOVA F-test and categorical variables with the chi-square test. Second, we compared the frequencies of patients who required ventilation, died or were discharged from the hospital by treatment status using the chi-square test. Third, to assess the association between treatment status and the study outcomes we estimated the Fine and Gray competing risk proportional hazards model.^12,13^ Models analyzing the outcome of death took into account the competing risk of discharge. Models analyzing the outcome of ventilation took into account the competing risks of discharge and death prior to ventilation. Using the Fine and Gray proportional hazards model we estimated the subdistribution hazard ratio, which represents the instantaneous event rate in patients who have not experienced the event or experienced a competing event.^12^ The proportional hazards assumption was tested as previously described^14^ using the implementation within the R package goftte.^15^ No violations of the proportional hazard assumption were identified. To account for non-randomized assignment to the treatment groups, we utilized propensity score adjustment. For the outcomes of death and death after ventilation, we created propensity scores for hydroxychloroquine use alone and hydroxychloroquine and azithromycin use during the hospital stay. For the ventilation outcome, we created propensity scores for hydroxychloroquine use alone prior to ventilation and hydroxychloroquine and azithromycin use prior to ventilation. Both sets of propensity scores were estimated via multinomial logistic regression of treatment group. All baseline covariates were included in the propensity score models. The propensity scores were entered into the outcome models with restricted cubic splines.^16^ Statistical analyses were performed with the use of SAS software, version 9.4 (SAS Institute) and R software, version 3.6.1 (the R project [http://r-project.org]).

## RESULTS

We identified all 385 patients who were hospitalized with confirmed SARS-CoV-2 infection at Veterans Health Administration medical centers across the United States and either died or were discharged as of April 11, 2020. Because the number of female patients in this cohort (17) was too small to permit robust statistical analyses, we evaluated the remaining 368 male patients in this study. Patients were categorized into three different groups: those treated with hydroxychloroquine (HC, n=97), treated with hydroxychloroquine and azithromycin (HC+AZ, n=113), or unexposed to hydroxychloroquine (no HC, n=158) (Table 1). All patients received standard supportive management. There were significant differences among the three groups in baseline demographic characteristics, selected vital signs, laboratory tests, prescription drug use, and comorbidities (Table 2).

**Table 1.** Categorization of patients based on pre-ventilation treatment.

|  | **HC** | **HC+AZ** | **No HC** | **Total** |
| --- | --- | --- | --- | --- |
| Treatment during all hospitalization period (N) | 97 | 113 | 158 | 368 |
| Treatment prior to ventilation (if ventilated) (N) | 90 | 101 | 177 | 368 |

HC: hydroxychloroquine-treated
HC+AZ: hydroxychloroquine and azithromycin-treated
No HC: not treated with hydroxychloroquine
A total of 368 patients met inclusion criteria and were assigned to one of three cohorts based on medication exposure: (1) HC-treated; (2) HC- and AZ-treated; or (3) HC-untreated. Additionally, to examine the association with ventilation (if ventilated), the time of hydroxychloroquine and azithromycin dispense was coded dynamically, before or after ventilator support.

**Table 2.**
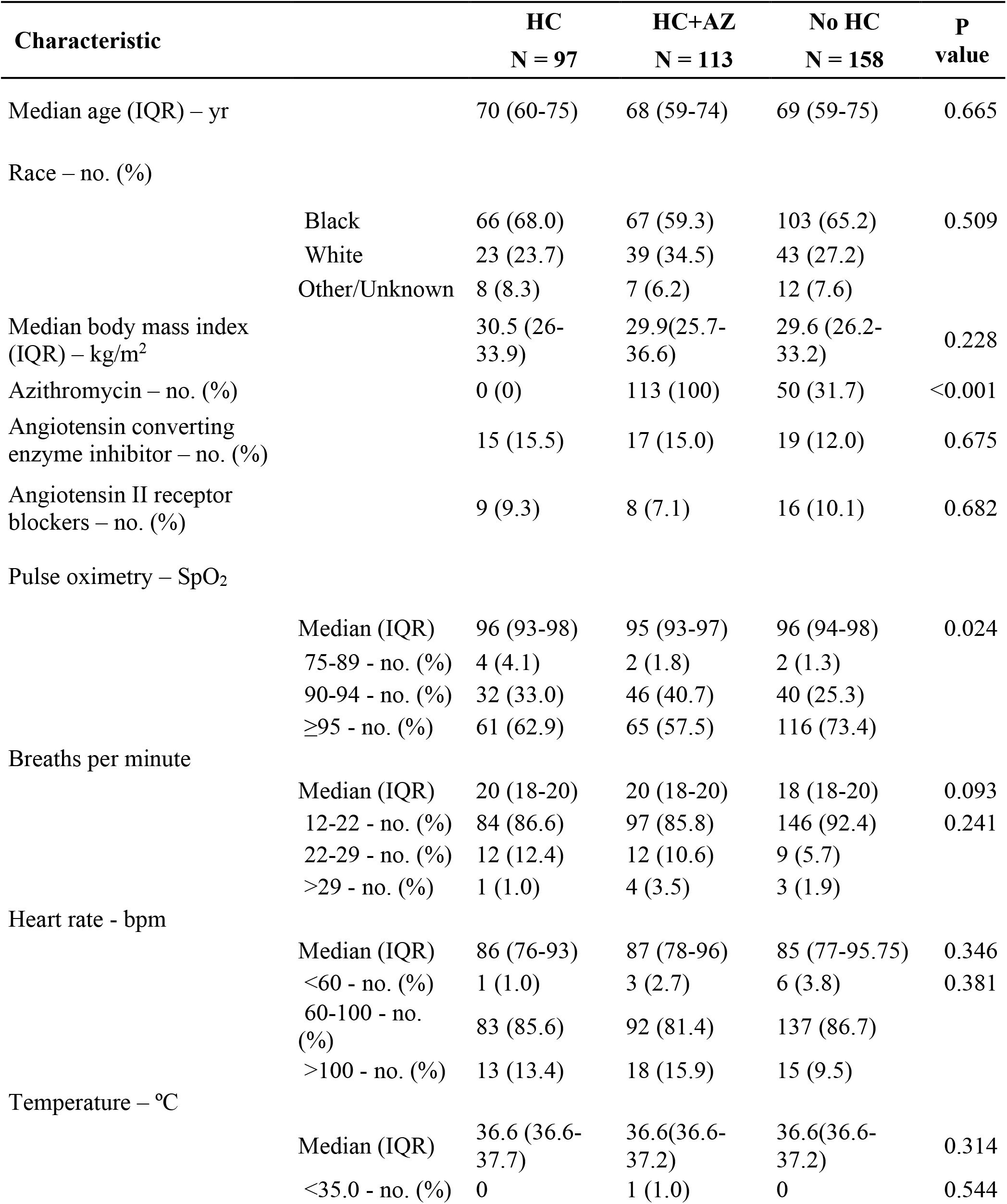
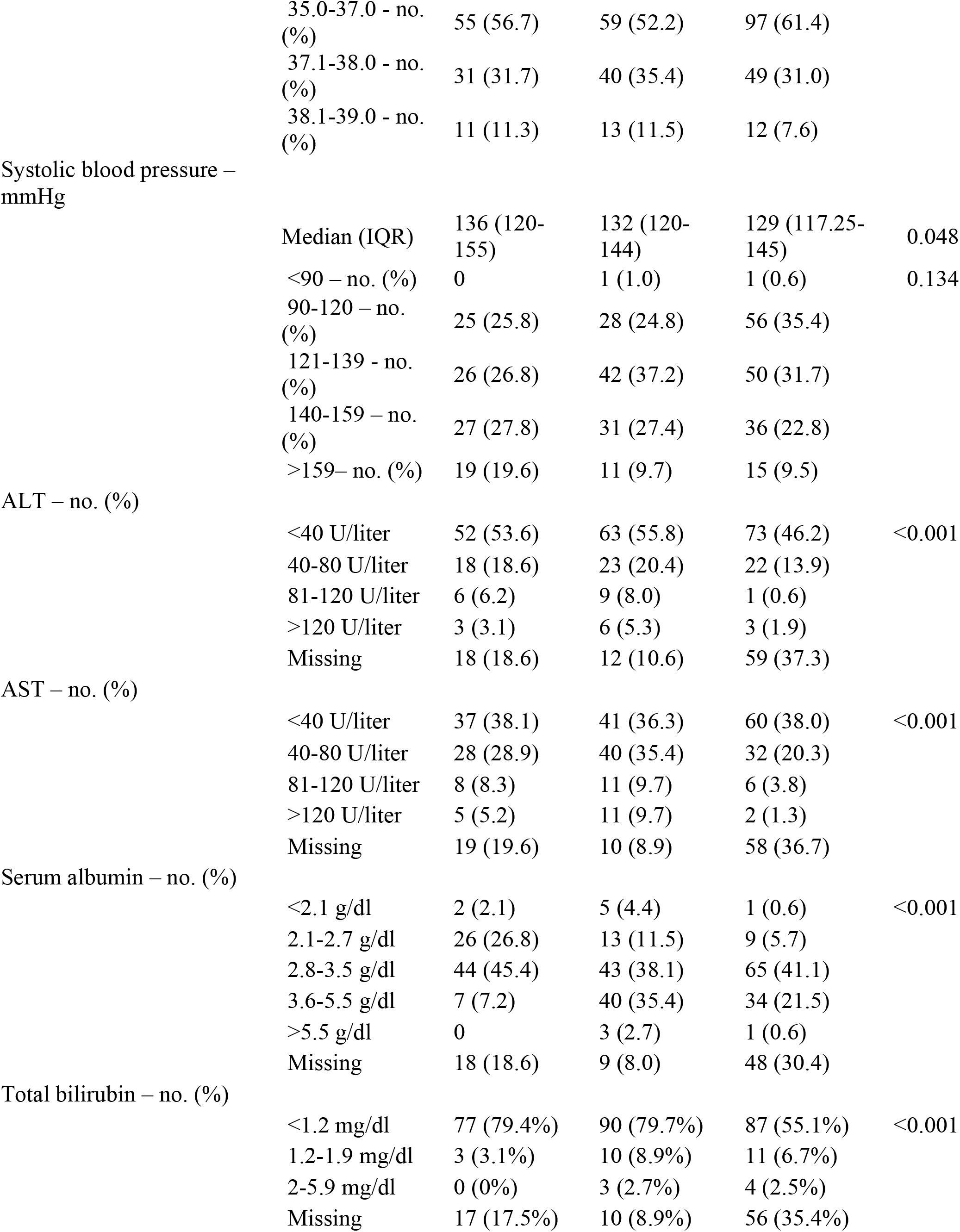
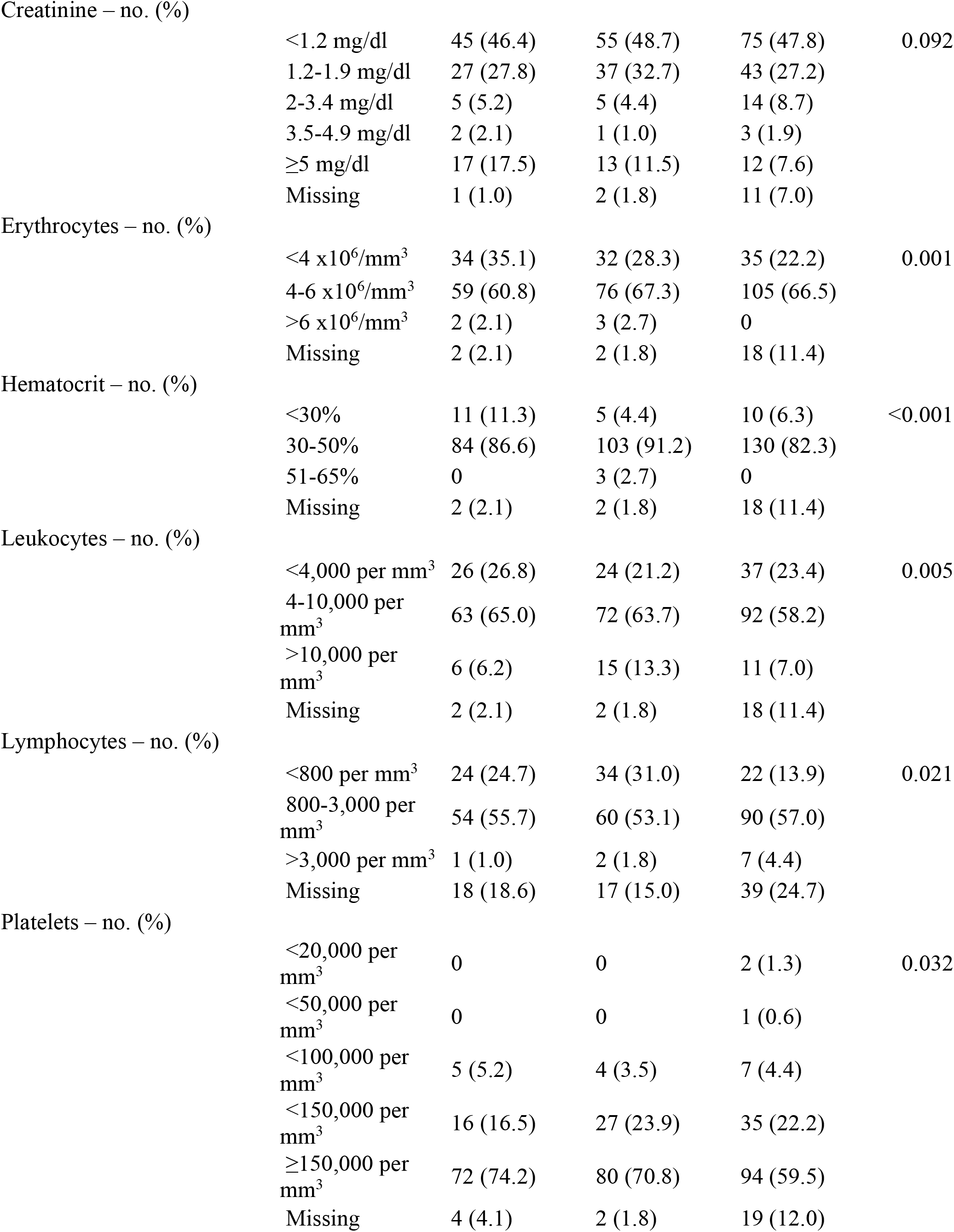
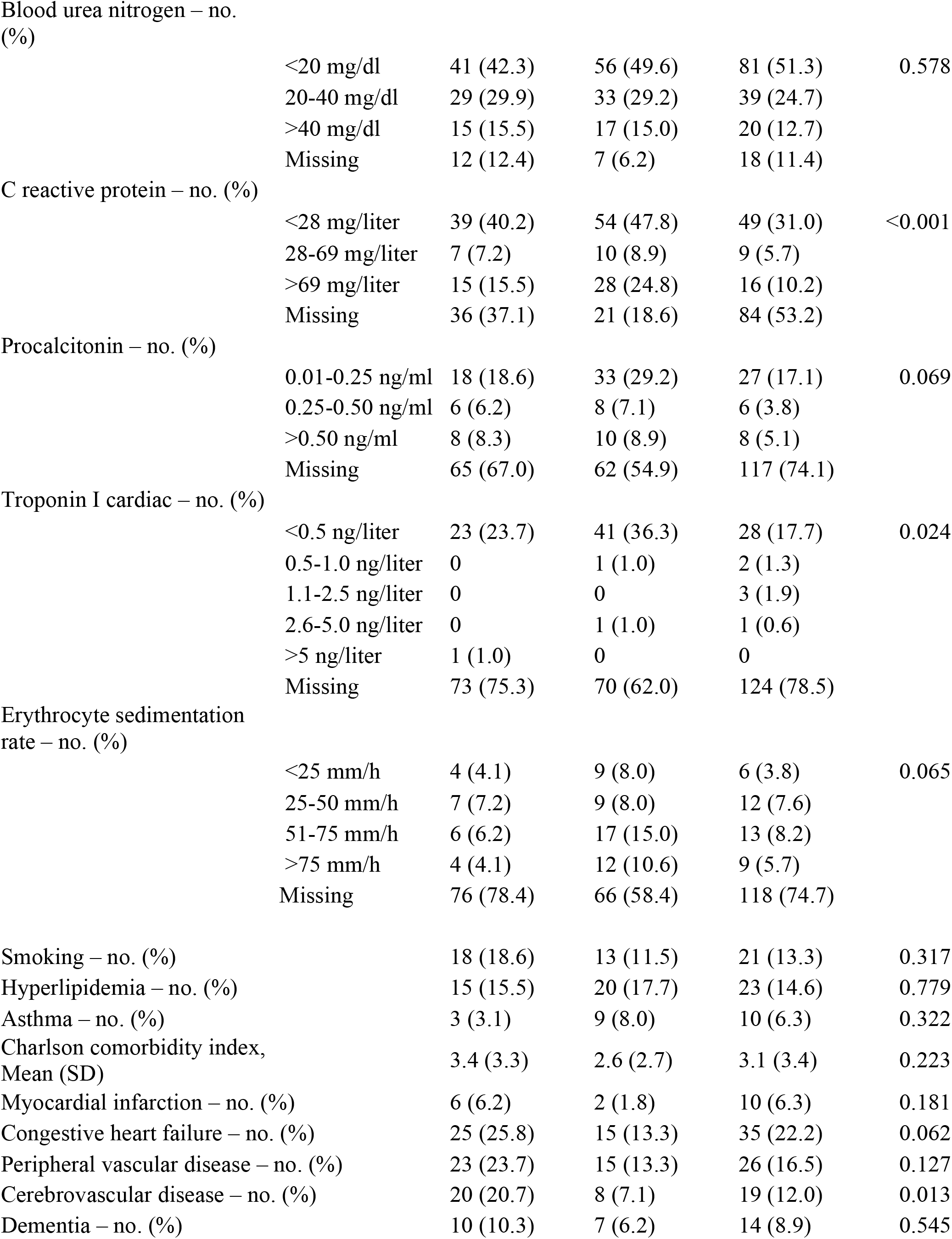
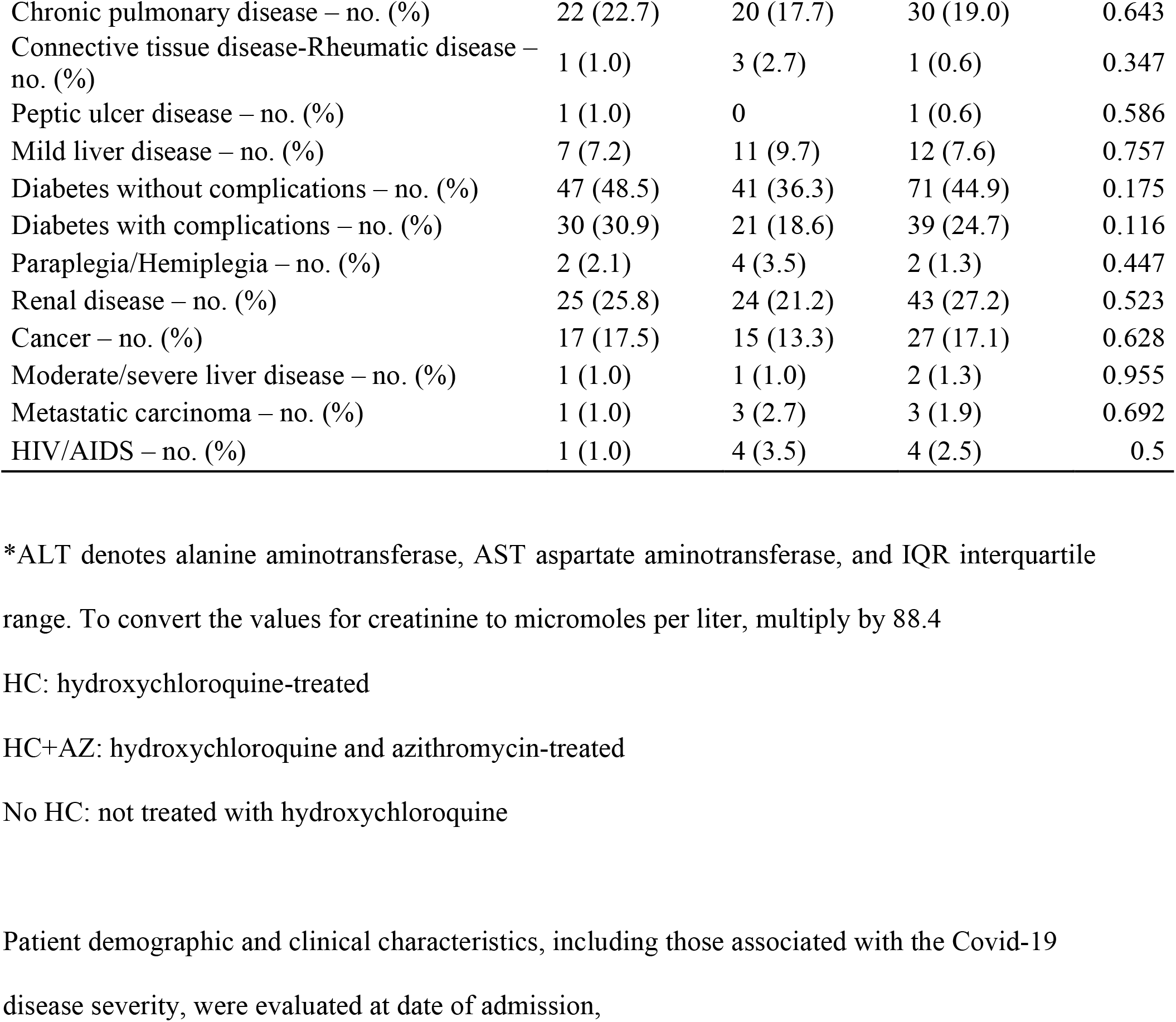
Baseline Demographic and Clinical Characteristics of the Patients.*

There were 27 deaths (27.8%) in the HC group, 25 deaths (22.1%) in the HC+AZ group, and 18 deaths (11.4%) in the no HC group (Table 3). Mechanical ventilation occurred in 13.3% of the HC group, 6.9% of the HC+AZ group, and 14.1% of the no HC group (Table 4).

**Table 3.** Outcomes based on treatment exposure.

| Outcome | HC<br>N=97 | HC+AZ<br>N=113 | No HC<br>N=158 | P value |
| --- | --- | --- | --- | --- |
| Death – no. (%) | 27 (27.8) | 25 (22.1) | 18 (11.4) | 0.003 |
| Discharge – no. (%) | 70 (72.2) | 88 (77.9) | 140 (88.6) |  |

HC: hydroxychloroquine-treated
HC+AZ: hydroxychloroquine and azithromycin-treated
No HC: not treated with hydroxychloroquine
Among the 368 patients evaluated, there were a total of 70 deaths. Patients not treated with hydroxychloroquine (No HC) had the lowest rate of death compared to the HC and HC+AZ cohorts.

**Table 4.**
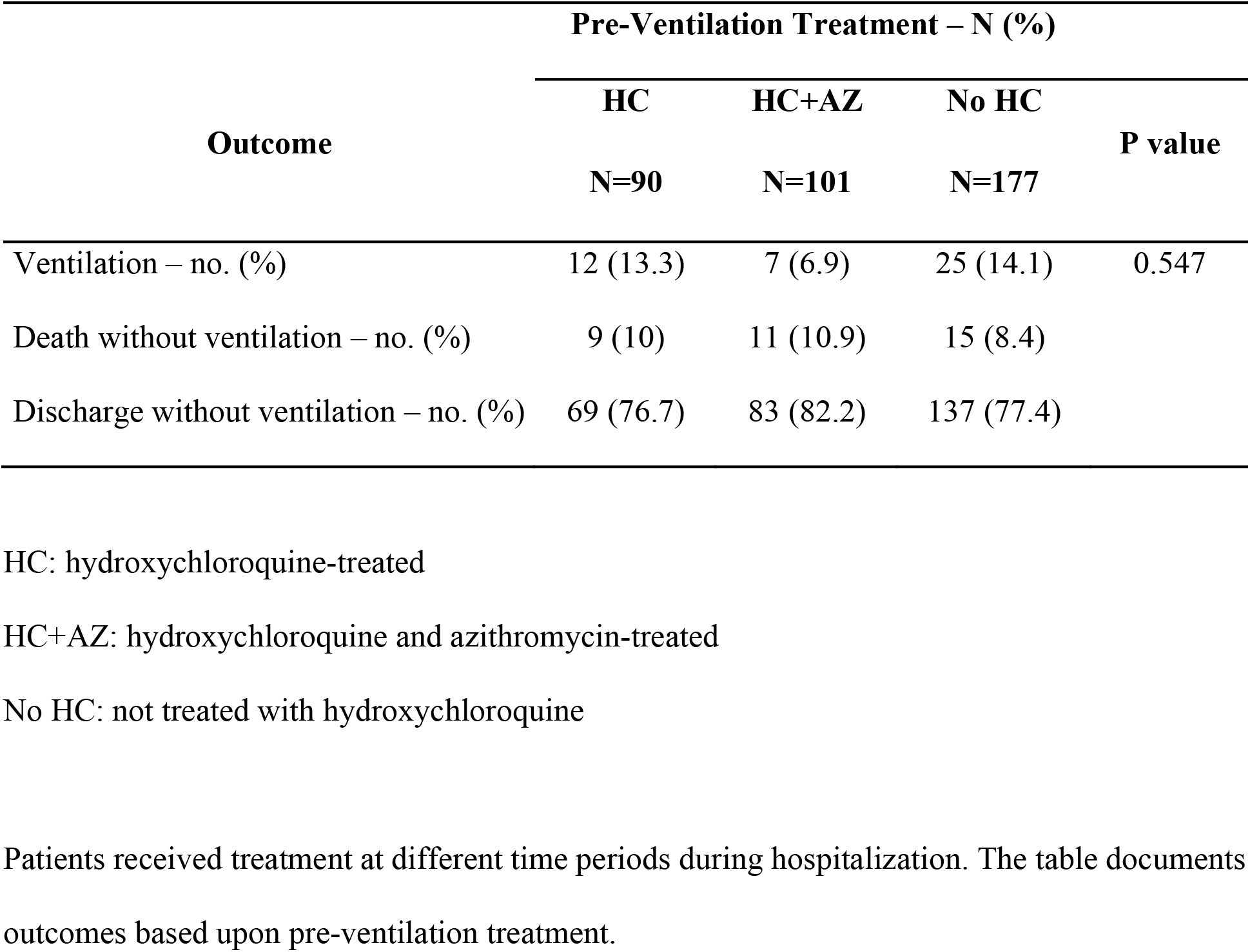
Outcomes based on pre-ventilation treatment.

| **Outcome** | **Pre-Ventilation Treatment – N (%)** |  |  | **P value** |
| --- | --- | --- | --- | --- |
|  | **HC** | **HC+AZ** | **No HC** |  |
|  | **N=90** | **N=101** | **N=177** |  |
| Ventilation – no. (%) | 12 (13.3) | 7 (6.9) | 25 (14.1) | 0.547 |
| Death without ventilation – no. (%) | 9 (10) | 11 (10.9) | 15 (8.4) |  |
| Discharge without ventilation – no. (%) | 69 (76.7) | 83 (82.2) | 137 (77.4) |  |

HC: hydroxychloroquine-treated
HC+AZ: hydroxychloroquine and azithromycin-treated
No HC: not treated with hydroxychloroquine
Patients received treatment at different time periods during hospitalization. The table documents outcomes based upon pre-ventilation treatment.

We analyzed the association of HC or HC+AZ use with the risk of overall death and the risk of ventilation as the primary outcomes. As baseline characteristics corresponding to clinical severity varied across the three groups of patients and could have influenced the non-randomized utilization of hydroxychloroquine and azithromycin, we computed propensity scores for HC use and HC+AZ use based on all baseline characteristics. Models analyzing the outcome of death took into account the competing risk of discharge. Models analyzing the outcome of ventilation took into account the competing risks of discharge and death prior to ventilation. The risks of these outcomes were estimated using subdistribution hazard regression.

Compared to the no HC group, there was a higher risk of death from any cause in the HC group (adjusted HR, 2.61; 95% CI, 1.10 to 6.17; P=0.03) but not in the HC+AZ group (adjusted HR, 1.14; 95% CI, 0.56 to 2.32; P=0.72) (Table 5). We did not observe a significant difference in the risk of ventilation in either the HC group (adjusted HR, 1.43; 95% CI, 0.53 to 3.79; P=0.48) or the HC+AZ group (adjusted HR, 0.43; 95% CI, 0.16 to 1.12; P=0.09), compared to the no HC group (Table 5).

**Table 5.** Subdistribution hazard of ventilation, death and death after ventilation.

|  |  | **Adjusted Hazard Ratio (95% Confidence Interval)** |  |  |
| --- | --- | --- | --- | --- |
|  |  | **Ventilation** | **Death** | **Death after ventilation** |
| HC | vs. No HC | 1.43 (0.53-3.79) | 2.61 (1.10-6.17) | 4.08 (0.77-21.70) |
| HC+AZ |  | 0.43 (0.16-1.12) | 1.14 (0.56-2.32) | 1.20 (0.25-5.77) |

HC: hydroxychloroquine-treated
HC+AZ: hydroxychloroquine and azithromycin-treated
No HC: not treated with hydroxychloroquine
The association between treatment and the outcomes using competing risk hazard regression adjusting for all clinical characteristics via propensity scores. Discharge and death were taken into account as competing risks and subdistribution hazard ratios are presented.

We then analyzed a secondary outcome of death among patients who required mechanical ventilation (Table 1). No significant difference was observed in the risk of death after ventilation in either the HC group (adjusted HR, 4.08; 95% CI, 0.77 to 21.70; P=0.10) or the HC+AZ group (adjusted HR, 1.20; 95% CI, 0.25 to 5.77; P=0.82), compared to the no HC group (Table 5).

## DISCUSSION

No effective therapy for Covid-19 has yet been identified. Given the longer development, testing, and approval times for novel chemical entities, repurposing drugs already approved for other indications is a promising approach to rapidly identify an effective therapy. Hydroxychloroquine is at the forefront of drug repurposing candidates Although ongoing randomized, controlled studies are expected to provide more informative evidence about hydroxychloroquine in the coming months, the outcomes observed in our study represent the best available data. This nationwide retrospective study of the largest integrated healthcare system in the United States provides the largest dataset yet reported of the outcomes of Covid-19 patients treated with hydroxychloroquine, with or without azithromycin, anywhere in the world. Specifically, hydroxychloroquine use with or without co-administration of azithromycin did not improve mortality or reduce the need for mechanical ventilation in hospitalized patients. On the contrary, hydroxychloroquine use alone was associated with an increased risk of mortality compared to standard care alone.

Baseline demographic and comorbidity characteristics were comparable across the three treatment groups. However, hydroxychloroquine, with or without azithromycin, was more likely to be prescribed to patients with more severe disease, as assessed by baseline ventilatory status and metabolic and hematologic parameters. Thus, as expected, increased mortality was observed in patients treated with hydroxychloroquine, both with and without azithromycin. Nevertheless, the increased risk of overall mortality in the hydroxychloroquine-only group persisted after adjusting for the propensity of being treated with the drug. That there was no increased risk of ventilation in the hydroxychloroquine-only group suggests that mortality in this group might be attributable to drug effects on or dysfunction in non-respiratory vital organ systems. Indeed, hydroxychloroquine use in Covid-19 patients has been associated with cardiac toxicity.^4^

Hydroxychloroquine has been reported to inhibit SARS-CoV-2 replication *in vitro* with a 50% maximal effective concentration (EC_50_) ranging from 4.5 μM to 17 μM.^17^ However, the approved dosing regimens for hydroxychloroquine in patients with rheumatoid arthritis or lupus generate substantially lower peak serum drug concentrations (∼1 μM).^18,19^ Administering higher doses of hydroxychloroquine to achieve presumed antiviral concentrations might increase the risk of adverse events. Interestingly, a randomized, controlled trial of high-dose chloroquine, the parent compound of hydroxychloroquine that also has been reported to have *in vitro* antiviral activity against SARS-CoV-2^17^ and similar peak serum concentrations in humans, was halted prematurely due to cardiac toxicity and higher fatality rates in the high-dose chloroquine-treated Covid-19 patients.^20^

Our study has certain limitations including those inherent to all retrospective analyses such as non-randomization of treatments. We did, however adjust for a large number of Covid-19-relevant confounders including comorbidities, medications, clinical and laboratory abnormalities. Despite propensity score adjustment for a large number of relevant confounders, we cannot rule out the possibility of selection bias or residual confounding. Our study cohort comprised only men whose median age was over 65 years. Therefore, the results may not necessarily reflect outcomes in women or in younger hospitalized populations, nor can they be extrapolated to pediatric patients. Our findings may also be influenced by the demographic composition of patients in our cohort, the majority of whom were black. Disproportionately higher rates of Covid-19-related hospitalization among the black population have also been reported in the United States as a whole.^21^

Our study also has certain strengths. Because we studied data from a comprehensive electronic medical record rather than from an administrative health insurance claims database, we used rigorously identified covariates and outcomes. We studied patients in an integrated national healthcare system; therefore, the data are less susceptible to biases of single-center or regional studies.

Data from ongoing, randomized controlled studies will prove informative when they emerge. Until then, the findings from this retrospective study suggest caution in using hydroxychloroquine in hospitalized Covid-19 patients, particularly when not combined with azithromycin.

## Data Availability

Data were extracted from the Veterans Affairs Informatics and Computing Infrastructure (VINCI), and thus are not publicly available.

## ACKNOWLEDGMENTS

This work was supported by National Institutes of Health (USA) grants (R01EY028027 and R01EY029799), DuPont Guerry, III, Professorship, and University of Virginia Strategic Investment Fund to JA. The funders had no role in study design or conduct, data collection, analysis, or interpretation, manuscript writing, or the decision to submit the manuscript for publication. The content of this article is solely the responsibility of the authors and does not necessarily represent the official views of the National Institutes of Health, U.S. Department of Veterans Affairs, nor does mention of trade names, commercial products, or organizations imply endorsement by the United States government. This paper represents original research conducted using data from the Department of Veterans Affairs and is, in part, the result of work supported with resources and the use of facilities at the Dorn Research Institute, Columbia VA Health Care System, Columbia, South Carolina.

JA is a co-founder of iVeena Holdings, iVeena Delivery Systems and Inflammasome Therapeutics, and has received consultancy fees from Allergan, Biogen, Boehringer Ingelheim, Immunovant, Janssen, Olix Pharmaceuticals, Retinal Solutions, and Saksin LifeSciences, all unrelated to this work. JA is named as an inventor on a patent application filed by the University of Virginia relating to Covid-19 but unrelated to this work. SSS has received research grants from Boehringer Ingelheim, Gilead Sciences, Portola Pharmaceuticals, and United Therapeutics, all unrelated to this work. The other authors declare no competing interests.

